# Large-Scale Application of Named Entity Recognition to Biomedicine and Epidemiology

**DOI:** 10.1101/2022.09.22.22280246

**Authors:** Shaina Raza, Deepak John Reji, Femi Shajan, Syed Raza Bashir

## Abstract

**Background:** Despite significant advancements in biomedical named entity recognition methods, the clinical application of these systems continues to face many challenges: (1) most of the methods are trained on a limited set of clinical entities; (2) these methods are heavily reliant on a large amount of data for both pretraining and prediction, making their use in production impractical; (3) they do not consider non-clinical entities, which are also related to patient’s health, such as social, economic or demographic factors.

**Methods:** In this paper, we develop Bio-Epidemiology-NER (https://pypi.org/project/Bio-Epidemiology-NER/) an open-source Python package for detecting biomedical named entities from the text. This approach is based on Transformer-based approach and trained on a dataset that is annotated with many named entities (medical, clinical, biomedical and epidemiological). This approach improves on previous efforts in three ways: (1) it recognizes many clinical entity types, such as medical risk factors, vital signs, drugs, and biological functions; (2) it is easily configurable, reusable and can scale up for training and inference; (3) it also considers non-clinical factors (age and gender, race and social history and so) that influence health outcomes. At a high level, it consists of the phases: preprocessing, data parsing, named entity recognition and named entities enhancement.

**Results:** Experimental results show that our pipeline outperforms other methods on three benchmark datasets with macro-and micro average F1 scores around 90 percent and above.

**Conclusion:** This package is made publicly available for use by researchers, doctors, clinicians and anyone to extract biomedical named entities from unstructured biomedical texts.

**Author Summary:** This paper introduces and presents a python package https://pypi.org/project/Bio-Epidemiology-NER/ that can extract named entities from the biomedical texts. Different from previous works, this package extracts not only clinical entities, such as disease, signs, symptoms but also demographics of the patients from the texts. This package can be used with least code requirements and can be used by epidemiologists, doctors, practitioners or others in the field to see the named entities from texts. The knowledge gained from the named entities help the end users to see the statistics or spread of infectious disease in least time and while parsing a large amount of free texts.

## 1 Introduction

Named entity recognition (NER) (1), a subtask of Natural Language Processing (NLP), seeks to identify and classify named entities (such as, person, place, event) in the unstructured text into pre-defined categories. In the biomedical domain, a fundamental task of NLP is the recognition of named entities, such as genes, diseases, species, chemicals, medical codes, drug names and so (2). NER can be used to extract important information from biomedical and clinical texts that can be used for many purposes, such as to study the statistical significance of certain entities (diseases, conditions), events, classification or relation extraction tasks (3). The recognition of clinical information through NER on a typical medical record is shown in Figure 1.

**Figure 1:**
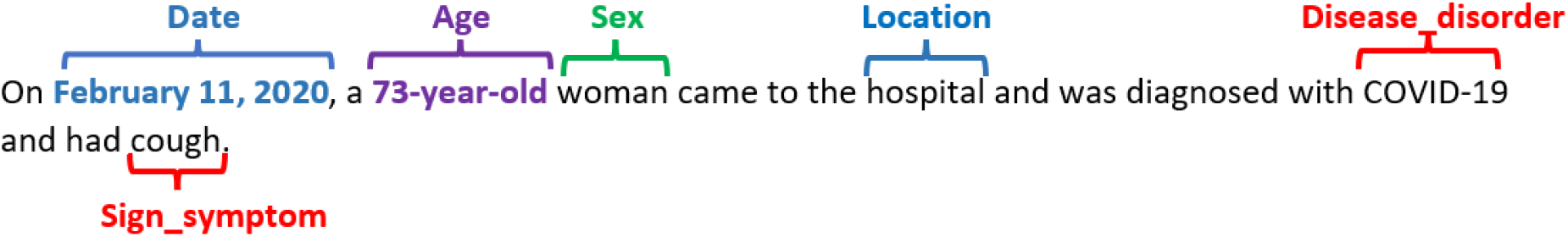
Example of the medical records NER result

As illustrated in Figure 1, it is possible to extract medical information from a typical medical record, such as disease disorder or signs/symptoms, as well as personal demographics (age, sex, location). The state-of-the-art work (4–6) in biomedical NER mostly focuses on limited named entities (disease, chemicals, genes, etc.). However, there are many biomedical entities that must be considered, particularly those related to clinical diagnosis, such as disease, symptoms, medical concepts, risk factors, vital signs; epidemiological entities such as infectious diseases, or patient demographics. This research is motivated by the need for an efficient and comprehensive biomedical NER that can automatically extract many entity types (clinical, epidemiological, demographics) from free texts (medical records, electronic health records, published literature) [7] in multiple formats (text, PDFs, rich text). The goal of this research is to facilitate the medical practitioners, clinicians, nurses and doctors in fast retrieval of information with a high degree of accuracy and efficiency.

In the state-of-the-art, the NER models are categorized into three main methods: early rule-based and dictionary-based methods (7), statistical machine-learning-based methods (8), and deep-learning based methods (2,9,10) in recent years. Although rule-based methods have demonstrated high accuracy, they require a large number of rules written by subject-matter experts, which is a limitation. Statistical machine learning methods also require an annotated corpus, which is not always feasible for large training tasks and limited resources (time and annotators). In recent years, the deep learning methods have demonstrated powerful generalization ability and has become the mainstream method for solving NER tasks (11). The development of hardware capabilities, emergence of distributed word representations and the availability of large training corpuses have contributed significantly to the success of deep neural network-based methods (12,13).

One of the most effective deep-neural network-based model is Bidirectional Encoder Representations from Transformers (BERT) (14), which is a multi-layer Transformer (13) model with self-attention (15). The original BERT model was trained on vast quantities of data for more than 104 languages, making its representations applicable to many smaller and similar downstream tasks, such as classification, NER, relation extraction. Research shows that the distillation of large language model can yield almost the same results as the original model, but we get the benefit of better efficiency and ease of use for production (16). In this research, we employ DistilBERT (17), a simplified version of the BERT with fewer parameters, faster training and better performance for the task of biomedical NER. Our contribution in this research is three-fold:

1. We develop a python package BioEN, short for **Bio**-**E**pidemiology-**N**er (https://pypi.org/project/Bio-Epidemiology-NER/), which can recognize accurate biomedical named entity annotations from the free text data (medical records, clinical notes, case reports, scientific publications). This package can parse text data in various input formats, including text files, tabular data, and PDF files. To facilitate analysis for end users, the model outputs named entities in both data frame, and annotated PDF formats (if the input is PDF file).
2. We make this package publicly available for distribution as software tools via PyPI and pip, making it easy for developers, researchers, and anyone with minimal programming knowledge to download and install it for both casual experiments and large, professional systems.
3. We provide a large number of biomedical entity types, such as diseases, risk factors, adverse events, and patient demographics, which is both extensive and more informative in comparison to previous works in this line of research.

Experimental results on several benchmark datasets showed the superiority of our approach compared to the state-of-the-art methods.

The rest of the paper is organized as: section 2 is the related work, section 3 is the methodology, section 4 is the experimental setup, section 5 is the results and analysis, section 6 is the discussion and section 7 is the conclusion.

## 2 Related work

Named entity recognition (NER) is the task of identifying a named entity (a real-world object or concept) in unstructured text and then classifying the entity into a standard category (1). These methods involve two tasks: (1) identification of entities (e.g., persons, organizations, locations, etc.) in text, and (2) classification of these entities into a set of predefined categories, such as person names, organizations (companies, government organizations, committees, etc.), locations (cities, countries, towns), date and time (1). Traditional NER methods only consider specific entities. However, there can be more entities, depending on the domain being used. For example, the field of biomedicine covers entities such as genes, diseases, chemicals, and proteins (2).

In recent years, there has been a dramatic increase in biomedical data (18). In particular, due to the COVID-19 surge, there has been a massive increase in biomedical data that is difficult to read, even more so when the urgency of time and the number of patients is increasing exponentially. To perform the biomedical mining, it is important to accommodate a prior process of biomedical NER. Biomedical NER is the task of identifying entities in the biomedical domain, such as chemical compounds, genes, proteins, viruses, disorders, drugs, adverse effects, metabolites, diseases, tissues, DNAs and RNAs, organs, toxins, food, or so (6,11). Most of the research (1,6,19) in biomedical NER focus on general approaches to named entities that are not specific to the biomedical field. On the other hand, there are some works (5,20–22) that focus solely on biomedical and chemical NER, however, they don’t cover many clinical entities. In this research, we plan to cover many named entities, that are both clinical, biomedical and epidemiological.

Word embedding is a useful technique that uses a large amount of unlabeled data to learn the latent syntactic and semantic information of words/tokens and map these words/tokens into dense low-dimensional vectors. In the past few years, many word embedding methods such Word2Vec [29], [30] and GloVe [31] are proposed. Nowadays, language models such as ELMo [15] and BERT [16] are considered as the state-of-the-art models. Unlike traditional word embeddings such as Word2Vec and GloVe, the embedding assigned to the word/token by the language model depends on the context, which means the same word/token could have different representations in different contexts. BERT [16] employs Transformer [32] to pre-train representations by jointly conditioning on both left and right context in all layers. Because the great success of BERT, it has gradually become a mainstream method using a large corpus to pre-train BERT and fine-tuning it on the target dataset. The BERT is also a widely used model now for many downstream tasks, such as NER and relation extractions.

Some biomedical works consider BERT for the biomedical NER tasks (5,23) and have shown outstanding performance. In this work, we also use the BERT model for NER task but we use the distilled version of the BERT, DistilBERT (17) in this work. The DistilBERT retains only half of the layers and the parameters of the actual BERT model. The distilled versions also balance between the computational complexity and the accuracy of the model, which is the motivation for our model building.

## 3 Materials and Methods

### 3.1 Problem definition

Given an input sentence *X* = {*x*_1_, *x*_2_, … ., *x*_*N*_}, where *x*_*i*_ is the *i*^th^ word (token) and *N* represents the length of the sentence. The goal of this study is to classify each token in *X* and assign it to a corresponding label *y* ∈ *Y*, where *Y* is a predefined list of all possible label types (e.g., disease, symptoms, drugs etc.).

Next, we present the workflow of BioEN development architecture in Figure 2.

**Figure 2:**
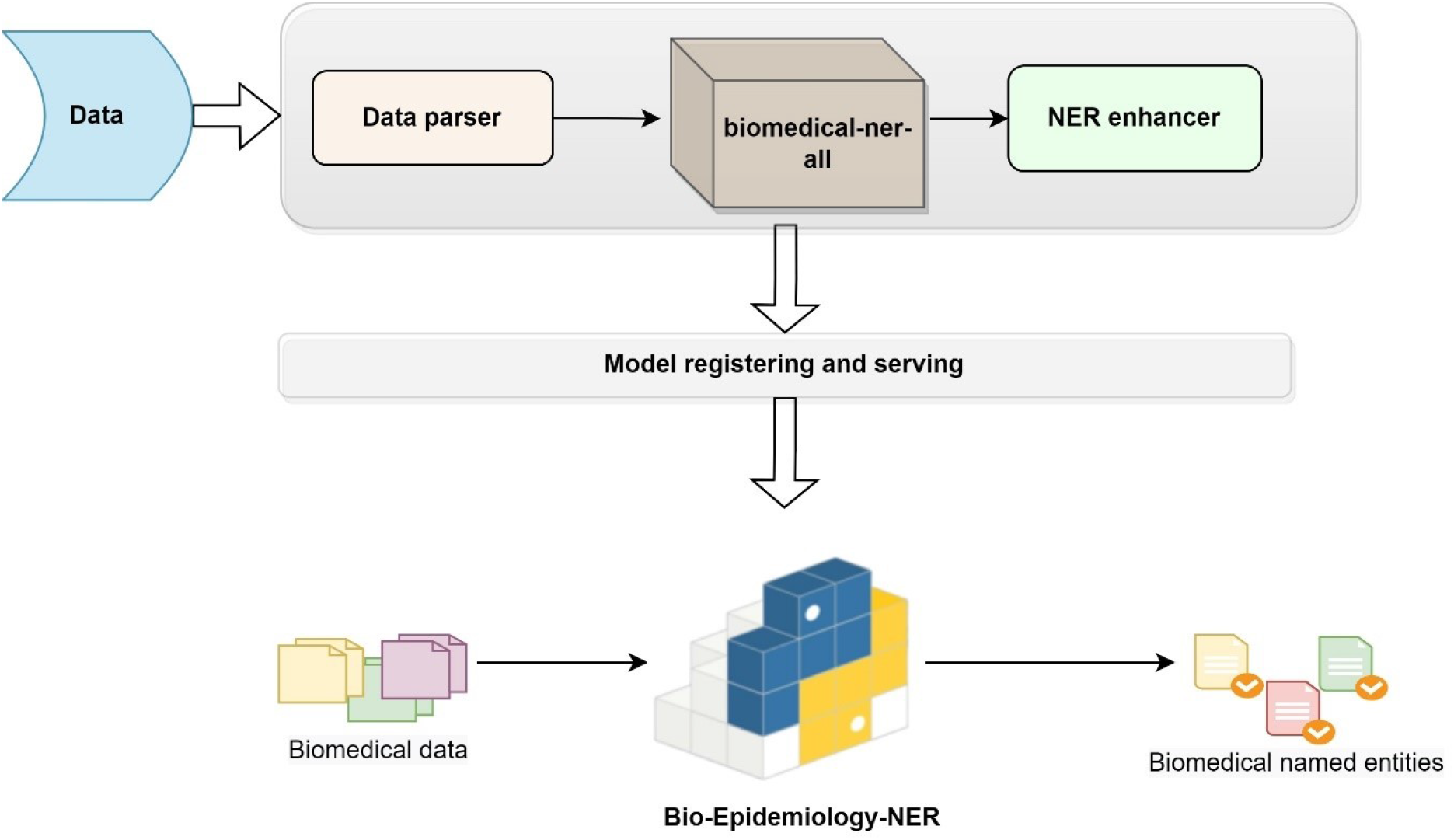
BioEN ((Bio-Epidemiology-NER) Development Architecture

### 3.2 Overall architecture

We have proposed and developed the BioEN package with a 3-phased approach in this study, as shown in Figure 2.

The details of each phase are given below:

First, we get the data, which is free texts, we feed the biomedical data in this architecture. In the first phase, which is the solution phase, (starting from the top) of the architecture, we build a fine-tuned Transformer model (we name it biomedical-ner-all model) with a data parser, and an NER enhancer. The second phase is the model registering and serving to prepare the models and related components for packaging. The third phase is the production phase, where we make our python package ready and deploy for real-time use. We explain the phases of BioEN architecture below:

#### 3.2.1 Solution phase

The first phase is the solution phase, where we have the data parser component, model processing and final output generation. We name this phase as ‘solution’ phase, since we are providing the main solutions to biomedical NER task here.

##### Data ingestion

The solution phase starts it working with the data ingestion. We can feed any free (unstructured) texts in any format (text files, rich texts, pdf files) to the data ingestion step. In this study, we feed the biomedical data (details in Section 4.1), however, we design this architecture with reusability in mind, so the same architecture can be used with any other domain specific data (e.g., life sciences, news feeds, entertainment).

##### Data parser

The free texts after data ingestion go into the *data parser* module. Data parsing is the process of converting data strings from one format to another, that is readable by the subsequent components in this phase. The data parser module can extract the textual data from any text file formats, including the PDF documents. The assumption behind such a multipurpose data parser is that the majority of biomedicine articles are available in PDF file formats.

##### Transformer-based model, biomedical-ner-all

The pre-processed texts from the data parser go into the next module, which is a Transformer-based module. We name this module *biomedical-ner-all* model and we make this module available at https://huggingface.co/d4data/biomedical-ner-all. The biomedical-ner-all model can identify the biomedical named entities from the texts. We have fine-tuned this model on biomedicine data that covers a large number of clinical, epidemiological and non-clinical (demographics) named entities. More details about biomedical-ner-all in Section 3.3.

##### NER enhancer

The output of the last module (i.e., Transformer-based model) is in the IOB (Inside-Outside-Before) format, which has become a prototypical standard format for tagging tokens in the NER tasks (24). The NER enhancer module is built on the top of biomnedical-ner-all model to convert the IOB representation to a user-friendly format by associating chunks (tokens of recognized named entities) with their respective labels and to enhance the NER predictions. We filter out the NER chunks with no associated entity (tagged ‘O’). The output of ner enchancer module are the named entities that are easily readable and are tagged. For example, we tag the exact token place for a disease mention, or a symptom or any other named entities as output representations.

#### 3.2.2 Model registering and serving

The second phase in the BioEN development architecture is the model registering and serving. In this phase, we bundle our ML models (in the first phase) into a package for local real-time inference as well as for batch inference. The idea is to support the real-time data processing and model serving workflow. We register the model and its components in a standard format for packaging (as in PyPi and pip) that can be used in a variety of downstream tools—for example, batch inference or real-time serving through a REST API. By the end of this phase, we have a package and a hosted model that can be called using a generalized code snippet, and we can load and run the model for the biomedical NER task.

#### 3.2.3 Biomedical-epidemiology-NER

The third phase in BioEN development architecture is the production phase. In this phase, we handle the versioning of our package and upload the final version to PyPI for distribution. The idea is to make this package available open-source, so that other developers, biostatisticians, epidemiologists, or researchers can easily download and install it either for casual experiments, or as part of large, professional systems. The package is available under MIT license with this link https://pypi.org/project/Bio-Epidemiology-NER/ and can be installed in any python environment by the following simple python command.

### pip install Bio-Epidemiology-NER

Below, we show the use of this package, which is quite simple, in Figure 3.

**Figure 3:**
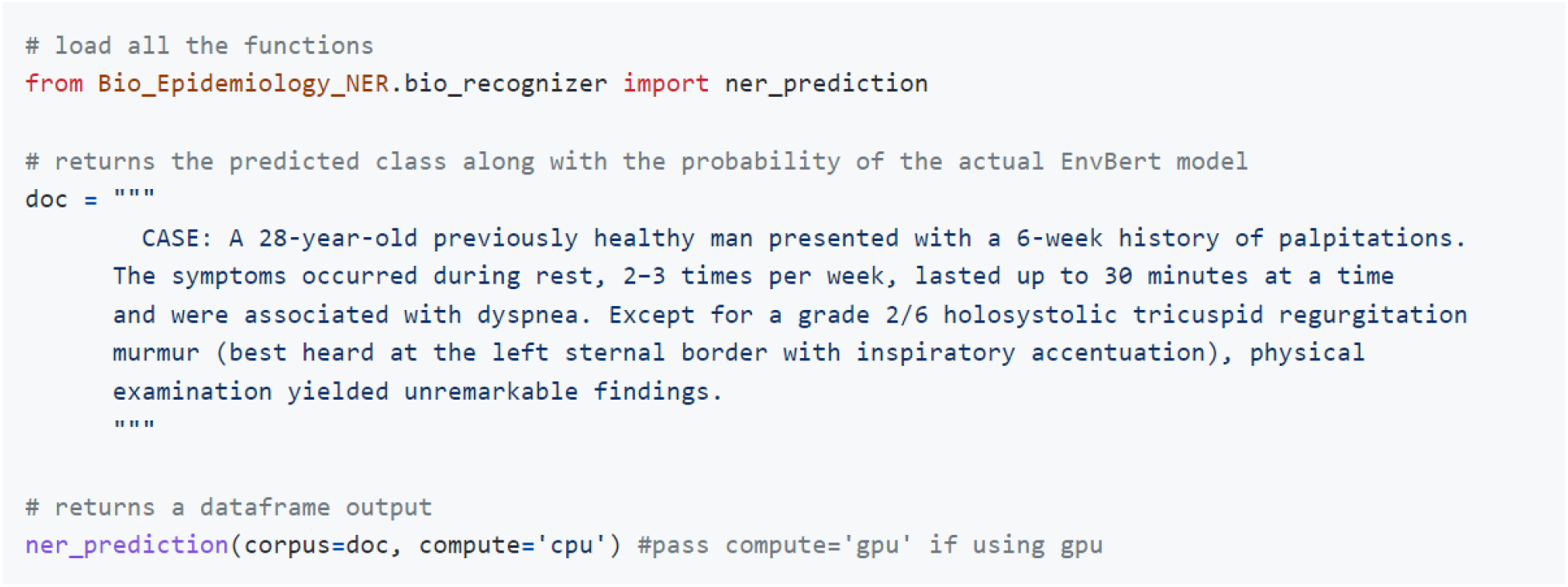
Illustration of package use on a case report.

The results are saved either in dataframes as CSV file with each chunk and named entity as output, or in the PDF files with entities being marked and shown. The tutorial and further details for use are given in the package website.

#### 3.2.4 Biomedical Named entities

The output of the package are biomedical named entities that are provided to the user in two formats: 1) data frames in CSV format with each word/ chunk and the identified named entity, and/or 2) pdf file with named entities highlighted and annotated in the texts.

In the state-of-the-art biomedicine works, there are usually a few named entities, such as disease, genes, proteins, chemical or species. However, we provide a large of number of clinical, non-clinical (demographics) and epidemiological named entities related to infectious diseases and the events, which we claim to be our unique contribution. We list the named entities that we are using in this work, in Table 1:

**Table 1:** Named entities used in this work

|  |  |  |
| --- | --- | --- |
| Activity | Diagnostic_procedure | Shape |
| Administration | Disease_disorder | Frequency |
| Age | Distance | Subject |
| Personal_background | Sign_symptom | Texture |
| Family_history | Medication | Therapeutic_procedure |
| Height | Outcome | Time |
| Sex | Severity | Volume |
| Color | Lab_value | Weight |
| Date | Mass | Dosage |
| Non-biological_location | History | Duration |
| Detailed_description | Coreference | Biological_attribute |
| Occupation | Qualitative_concept | Biological_structure |
| Personal- Biological_structure | Quantitative_concept | Clinical_event |
|  | Area | Other_entity |

Next, we explain our methodology, where we discuss our Transformer-based model ***biomedical-ner-all*** in detail.

### 3.3 Model details

BERT and its refined version, DistilBERT (17), are Transformer-based models with self-attention blocks. These models are pre-trained on unlabeled raw texts and can be used for a variety of tasks, including question answering, sentence-pair classification, and sequence tagging tasks (25). The idea is to have a general architecture that is applicable to many problems and a pre-trained model that reduces the need for labelled data. In this paper, we fine-tune the pre-trained DistilBERT model to learn a more accurate representation of target domain entities (biomedicine entities). We show the model adaption from a teacher model (pre-trained DistilBERT) to our student model (fine-tuned DistilBERT) in Figure 4.

**Figure 4:**
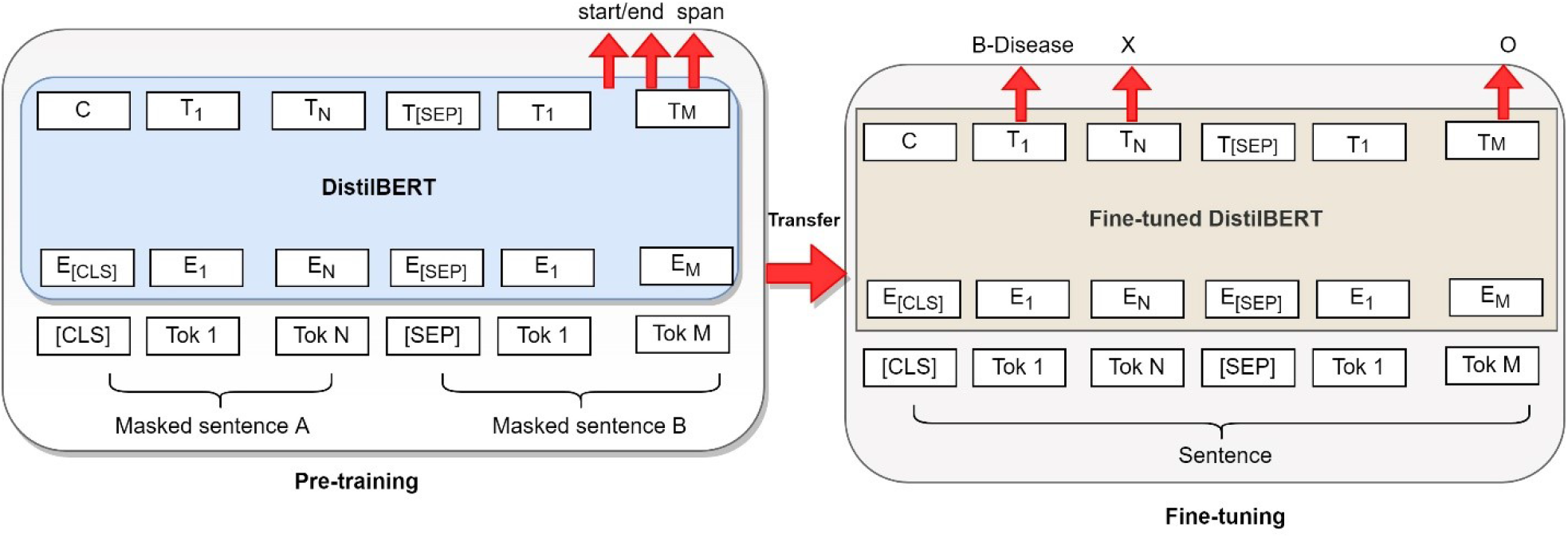
Model adaption from the teacher model to student model.

As shown in Figure 4, we fine-tine the pre-trained DistilBERT model’s weights for the initialization of the NER task, and we adjust the input and output to our biomedical task. We release the model weights of our fine-tuned model here^**Error! Bookmark not defined**.^. The model weights are also packaged in our python package^**Error! Bookmark not defined**..^

In the fine-tuning stage, we replace the entity tagging head of DistilBERT with a randomly initialized new head that covers all the entity categories of the target domain. We fine-tune the model using the target domain (biomedicine) training data. The model is optimized with Adam (26) with a batch size of 16 and a learning rate of 2e-5. We train the model for 40 epochs and evaluate the model on the development set using entity-level F1-score on sub-word tokens. In addition to early stopping, the model is regularized with dropout within each transformer block and weight decay. The dropout is set to 0.1, and the weight decay is set to be 0.01 in the experiments. The best performing model weights is used as the final prediction model.

As the input for the BERT model, we use the CoNLL-2003 (27) formatted data, which is split into sentences and tokenized on word-level. Sentences longer than the limit (512 sequence) are split into separate input sequences for the network. When converting the predictions back to the word-level CoNLL format, we assign the predicted entity label of the first sub-word unit for the entire token. Our fine-tuned model, which we name as ‘biomedical-ner-all’ released with our package also, is shown in Figure 5.

**Figure 5:**
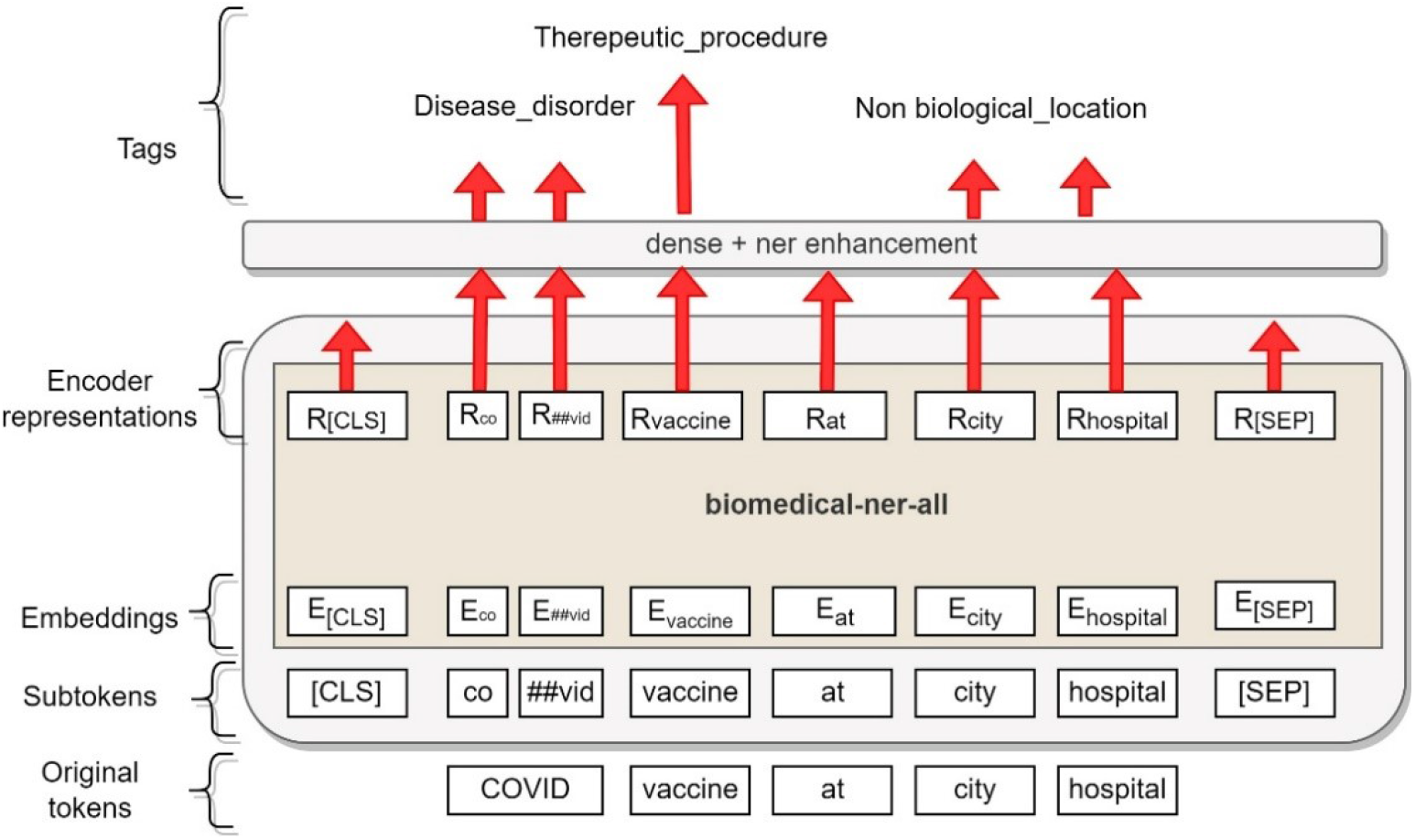
biomedical-ner-all model

As shown in Figure 5, we give the input, which are sequences of texts to the model. The input is tokenized and for each given token, its input representation is constructed by summing the corresponding token, segment, and position embeddings. The embedding of words goes as input to the dense layer, where we perform the enhancements to convert the IOB format of CONLL-2003 to a user-friendly format. The output is the set of entities that are user-friendly and easily readable. Along with extracting each entity, our model also shows a confidence score. The confidence score is simply a decimal number between 0 and 1 and it serves as an indicator of how confident the system is with its prediction. We consider only the entities which have prediction confidence score more than 0.4 in the final output.

### 3.4 Data

In this work, we use the publicly available data MACROBBAT 2020 dataset (28). The dataset details are available in the original paper (29), where the authors define the acronym ACROBBAT as ‘Annotation for Case Reports using Open Biomedical Annotation Terms’. The dataset consists of 200 source documents in plain text and 200 annotation documents, each annotation document with the plain text. Each document is named using PubMed document identifier, for example, 18258107.txt and 18258107.ann. The authors extracted the document’s text from the PubMed article, but only included the clinical case report information, which is also relevant to our application because it contains patients’ data.

As mentioned in the dataset paper (29), the documents were manually annotated by researchers who had prior experience reading biomedical and clinical language. Following completion, the annotations were checked for format and type consistency. We fine-tune our Transformer-based model (shown in Figure 5) with the MACROBBAT data, which covers a wide range of named entities (clinical, demographics, epidemiological and event-based). We also update the package with additional training on a portion of COVID-19 data, so that it can accurately detect the mentions of latest Coronavirus and/or COVID-19 named entities. The details of the dataset used in this study to train our model are given in Table 2:

**Table 2:** Dataset details

|  |  |
| --- | --- |
| **No. of documents** | 200 PubMed documents |
| **No. of sentences** | 3,652 |
| **No. of annotations** | 59,164 |
| **Main Entity types** | Clinical events, Diagnostic, procedure, disease disorder, medication, sign symptoms, time expressions, demographics, patient history, temporal and causal relations, co-references. |
| **Distinct entity types** | 42 (given in Table 1) |

Besides MACROBBAT, we also evaluate our approach against the following benchmark datasets:

- **NCBI-Disease** (30): it is the dataset introduced for disease NER and normalization. It has been widely used for a lot of applications. It consists of 793 PubMed abstracts with disease mentions.
- **I2b2-2012** (31): it consists of clinical (problems, tests, treatments, clinical departments), occurrences (admission, discharge and evidence) mentions using 310 discharge summaries.

We also collected random case reports (around 500) from the LitCOVID (32) data source from the year 2020-2021 to see the effectiveness of our approach on the biomedical and COVID-19 patients data.

### 3.5 Benchmarking methods and evaluation

#### Benchmarking

To assess the utility and challenges of our approach, we evaluate the performance of our NER approach against the following state-of-the-art models:

- **BiLSTM-CNN-Char** (33), a hybrid Bidirectional Long Short-Term Memory (LSTM) and Convolutional Neural Network (CNN) architecture that learns both character and word-level features for the NER task.
- **SciBERT** (Base), a pretrained language model based on BERT pretrained on a large multi-domain corpus of scientific publications to improve performance on downstream scientific NLP tasks.
- **BlueBERT** (34), BERT-based uncased model pretrained on PubMed abstracts and MIMIC-III.
- **ClinicalBERT** (35), BERT-Base-cased model trained on MIMIC notes.
- **BioBERT** (36), a pre-trained biomedical language representation model for biomedical text mining. We use the BioBERT-Base v1.2 (+ PubMed 1M).

These models have obtained state-of-the-art performance in their works, respectively.

#### Evaluation metrics

Following the standard practice (2,27,37) to evaluate NER tasks, we use the typically evaluation method, i.e., precision, recall and F1-score at a token level.

- Precision: percentage of named entities found by the learning system that are correct.
- Recall: percentage of named entities present in the data found by the system. A named entity is correct only if it is an exact match of the corresponding entity in the data file.
- F1-score: harmonic mean of precision and recall.

#### Configurations and Hyperparameters

We use PyTorch for the model implementation. We run our experiments on GPU: 1 x GeForce RTX 3060 with 16.0 GB RAM to integrate the components of our package. We use Grid search to get the optimal values for the hyperparameters and early stopping to overcome possible overfitting. We specify the following hyperparameters as shown in Table 3.

**Table 3.** Hyperparameters used

| Hyperparameter | Optimal value used |
| --- | --- |
| Learning rate | 2E-5 |
| Batch size | 16 |
| Epochs | 40 |
| Optimizer | Adam |
| Dropout rate | 0.5 |
| Optimizer | Adam |
| Hidden Size | 768 |
| Embedding Size | 128 |
| Max Seq Length | 512 |
| Warmup Steps | 3000 |
| Weight decay | 0.01 |
| dropout | 0.1 |

We have divided the dataset into training, validation, and test sets, with a 70:15:15 ratio for all experiments.

## 4 Results

In this section, we present the results and analysis.

### 4.1 Overall performance evaluation

We present the results of our model and the baseline models on all the datasets in Table 4.

**Table 4.** Overall performance

| Method | Metric | MACCROBAT | NCBI-Disease | I2b2-2012 |
| --- | --- | --- | --- | --- |
| BiLSTM-CNN-Char | Precision | 84.43 | 85.24 | 79.35 |
|  | Recall | 83.97 | 83.31 | 78.11 |
|  | F1-score | 84.20 | 84.26 | 78.73 |
| SciBERT | Precision | 78.10 | 76.88 | 77.01 |
|  | Recall | 72.18 | 74.10 | 75.18 |
|  | F1-score | 75.02 | 75.46 | 76.08 |
| BlueBERT | Precision | 84.04 | 83.37 | 81.10 |
|  | Recall | 81.48 | 81.39 | 80.88 |
|  | F1-score | 82.74 | 82.37 | 80.99 |
| ClinicalBERT | Precision | 81.01 | 84.08 | 80.35 |
|  | Recall | 79.10 | 80.11 | 78.69 |
|  | F1-score | 80.04 | 82.05 | 79.51 |
| BioBERT v1.2 | Precision | 86.72 | 85.80 | 88.00 |
|  | Recall | 88.31 | 84.29 | 86.10 |
|  | F1-score | 87.51 | 85.04 | 87.04 |
| **BioEN (our approach)** | Precision | **92.10** | **91.68** | **90.10** |
|  | Recall | **91.68** | **88.92** | **88.98** |
|  | F1-score | **91.89** | **90.28** | **89.54** |

Table 4 shows that our BioEN approach achieves the state-of-the-art performance for detecting a large number of biomedical named entities. This is demonstrated by our model outperforming other methods by achieving around 90% F1-score on all datasets. The superiority of our model is credit to its architecture and its carefully fine-tuned biomedical-ner-all model. This result also demonstrates that a model that is trained on domain-specific data (e.g., biomedical data) can be applied to a wide variety of domain-specific terminologies. For example, we train this package BioEN (including its components) on biomedical named entities and we are able to detect a large variety of clinical, event-based and epidemiological named entities to study the infectious diseases and population groups. Our model also considers the causal relations and co-references, which are provided by the MACCROBAT dataset on which it is primarily trained. Thus, this package is able to incorporate a diverse vocabulary and phenomena described in clinical documents without requiring direct connections to curated concepts (e.g., MeSH, UMLS knowledge bases).

We also see that BERT-based methods (BioBERT, BlueBERT and ClinicalBERT) that have been pre-trained on PubMed and/or MIMIC-III clinical notes, perform well. This is most likely due to the fact that these methods have been well-trained for extracting richer features, resulting in better overall performance. BioBERT outperforms other BlueBERT, ClinicalBERT and SciBERT, which is probably because BioBERT is pre-trained on more biomedical and clinical data and is able to better infer the patterns from the test data. We also find that BlueBERT performance is quite good, above 80%, which shows models pretrained on domain-specific literature (e.g., biomedicine) perform well in the respective downstream tasks.

As demonstrated in Table 4, BiLSTM-CNN-Char that automatically detects word- and character-level features using a hybrid BiLSTM and CNN architecture yields quite good result, after BioEN (ours) and BioBERT methods. BiLSTM-CNN-Char model achieves a F1-score performance of approximately 84% on MACCROBAT and NCBI dataset, which are quite high compared to most of the models. BiLSTM-CNN-Char has also been used as the state-of-the-art and a conventional model for many biomedical datasets. However, the latter works show that adding the language models on the top of conventional models (e.g., BiLSTM) show good performance (38,39), which may be attributed to large scale pre-training tasks on the top.

SciBERT is trained on papers from the semanticscholar.org corpus; while the knowledge gained is significant, the performance of the model in our work is somewhat compromised. We anticipate this comparatively lower performance may be a result of the limited biomedical literature used to train the model, and the named entity types of the actual model differ in some way from what is expected by our application.

Despite the fact that we fine-tune each baseline method to its optimal hyperparameter settings, we anticipate that the relatively low scores of these baselines on the MACCROBAT dataset, which is a primary dataset for our model training, can be because of few factors, for example, lack of complete training, and unavailability of the training/test set splits utilized in previous studies.

### 4.2 Effectiveness of the BioEN on case reports

We give a snippet from a COVID-19 related case report (40) to BioEN (our method) and show the confidence score for the predicted entities. Due to brevity reasons, we show the results only on one sentence from the case report. The results are shown in Table 5.

**Table 5.** Confidence score of the model on different named entities by BioEN

| Entity group | Value | Score |
| --- | --- | --- |
| Sex | man | 0.999528 |
| History | 40s | 0.867149 |
| Clinical_event | admitted | 0.999799 |
| Location | local hospital | 0.997007 |
| Date | 6 days after | 0.991963 |
| Disease_disorder | COVID-19 | 1 |
| Diagnostic_procedure | general condition | 0.999905 |
| Therapeutic_procedure | treated | 0.999675 |
| Location | intensive care unit | 0.999643 |
| Sign_symptom | cough | 0.999682 |
| Sign_symptom | experienced dyspnoea recurred and rapidly increased | 0.999941 |
| Sign_symptom | dyspnoea | 0.999938 |
| Sign_symptom | recurred | 0.992294 |
| Diagnostic_procedure | CT | 0.999842 |
| Biological_structure | pulmonary | 0.999952 |
| Diagnostic_procedure | angiogram | 0.999405 |
| Area | 10×18 cm | 0.997637 |
| Detailed_description | cavitary | 0.999947 |
| Detailed_description | pulmonary cavitary | 0.999942 |
| Detailed_description | air-fluid level | 0.999005 |
| Disease_disorder | atelectasis | 0.99756 |
| Detailed_description | one-way valve mechanism | 0.84776 |
| Coreference | developed pneumatocele | 0.863188 |
| Coreference | one-way pneumatocele | 0.792779 |
| Biological_structure | bronchial endobronchial segments | 0.915637 |
| Biological_structure | right lower lobe | 0.999896 |
| Biological_structure | endobronchial | 0.999509 |
| Date | 4 weeks after | 0.999853 |
| Date | Six months after | 0.999874 |
| Biological_structure | lung | 0.980932 |

As can be seen, our model is able to predict the named entities with quite accuracy, which is also validated by two experts in the biomedicine field.

### 4.3 Case study

We show the most frequent top-10 named entity types predicted by our model, through parsing 100 random case reports from PubMed, in Table 6. These case reports are selected from the timeline 2020-2021 and are related to COVID-19. These results show the predictions from our model.

**Table 6.** Most frequent named entities from 100 covid-19 case reports

| Disease disorder | Sign symptom | Therapeutic procedure | Medication |
| --- | --- | --- | --- |
| covid-19 | cough | arterial | aspirin |
| homocystinuria | fever | Endomyocardial | homocysteine |
| influenza | shortness of breath | intubation | alcohol-based |
| asthma | sore throat | oxygenation | Paxlovid |
| SARS | headache | splenectomy | Tylenol |
| gastroenteritis | aches and pains | vaccination | ibuprofen |
| long-COVID | diarrhoea | venoarterial | clopidogrel |
| cardiac | chest pain | Myocardial | prasugrel |
| respiratory | rash | Endovascular | acetaminophen |
| pneumonia | difficulty breathing | transplantation | antiviral pills |

These results in Table 6 can be used to get some valuable information regarding the most frequent disorders or symptoms mentioned in the case report or to find the most common findings in an efficient manner. According to these resuts, the most common symptom is cough and fever, while the most common drug ingredients mentioned is aspirin. We also see that most common disease disorder here is COVID-19, since these are COVID-19 case reports.

We also show the most common disease disorder in two sexes (male and female), based on 100 case reports data, and the results are shown in Figure 6.

**Figure 6:**
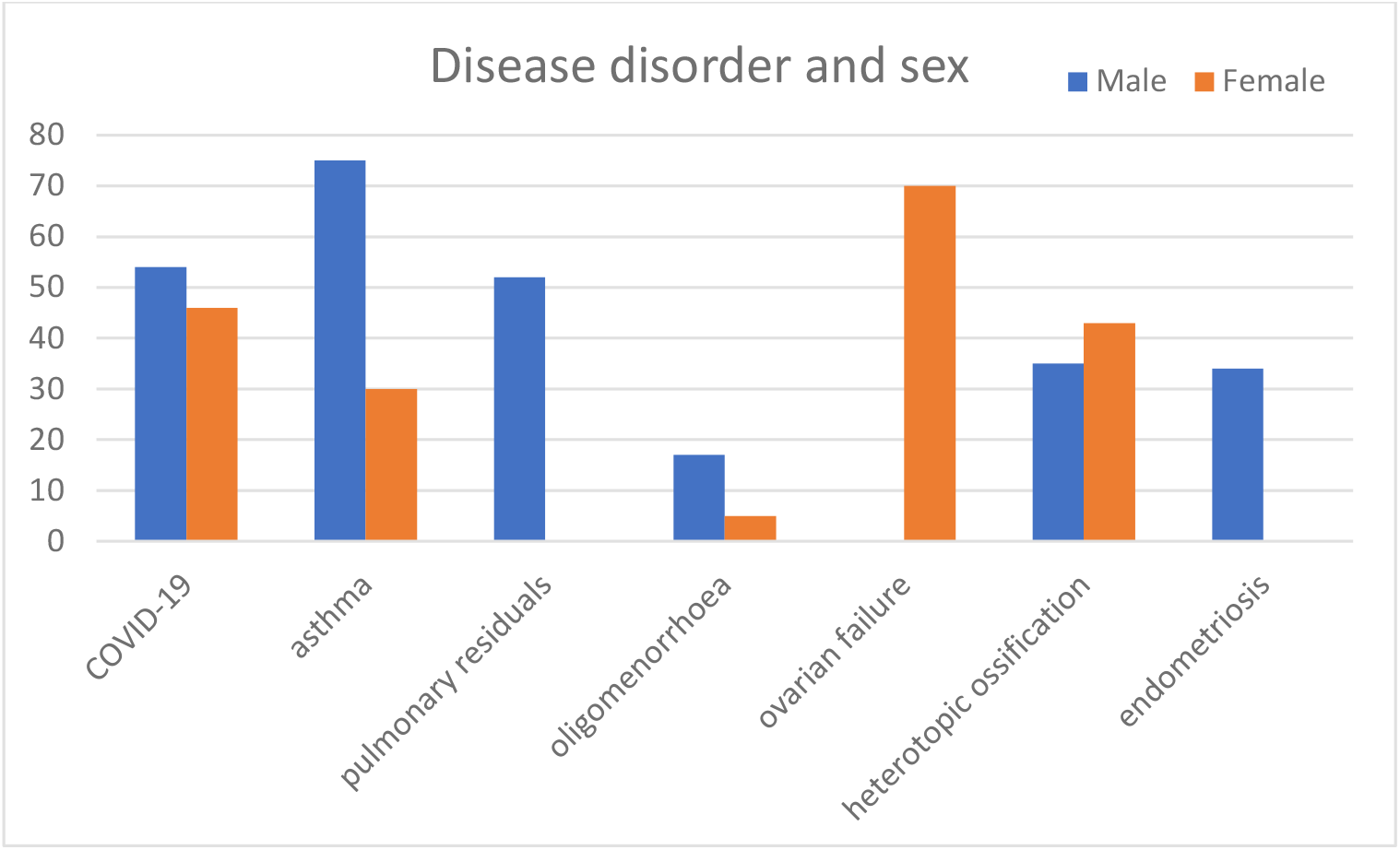
Distribution of disease disorders among male and female groups.

As shown in figure 6, COVID-19 patients around 54% as male and the other are females. Ovarian failure, which is a female disease order, is found is female patients only. For the other diseases, we see asthma mostly in male patients and endometrisis only in male patients.

We also show the percentage of most occurring diseases in the patients in Figure 7 and the most frequent diseases are COVID-19 positive, pneumonia, respiratory, which are primarily related to COVID-19 disease.

**Figure 7:**
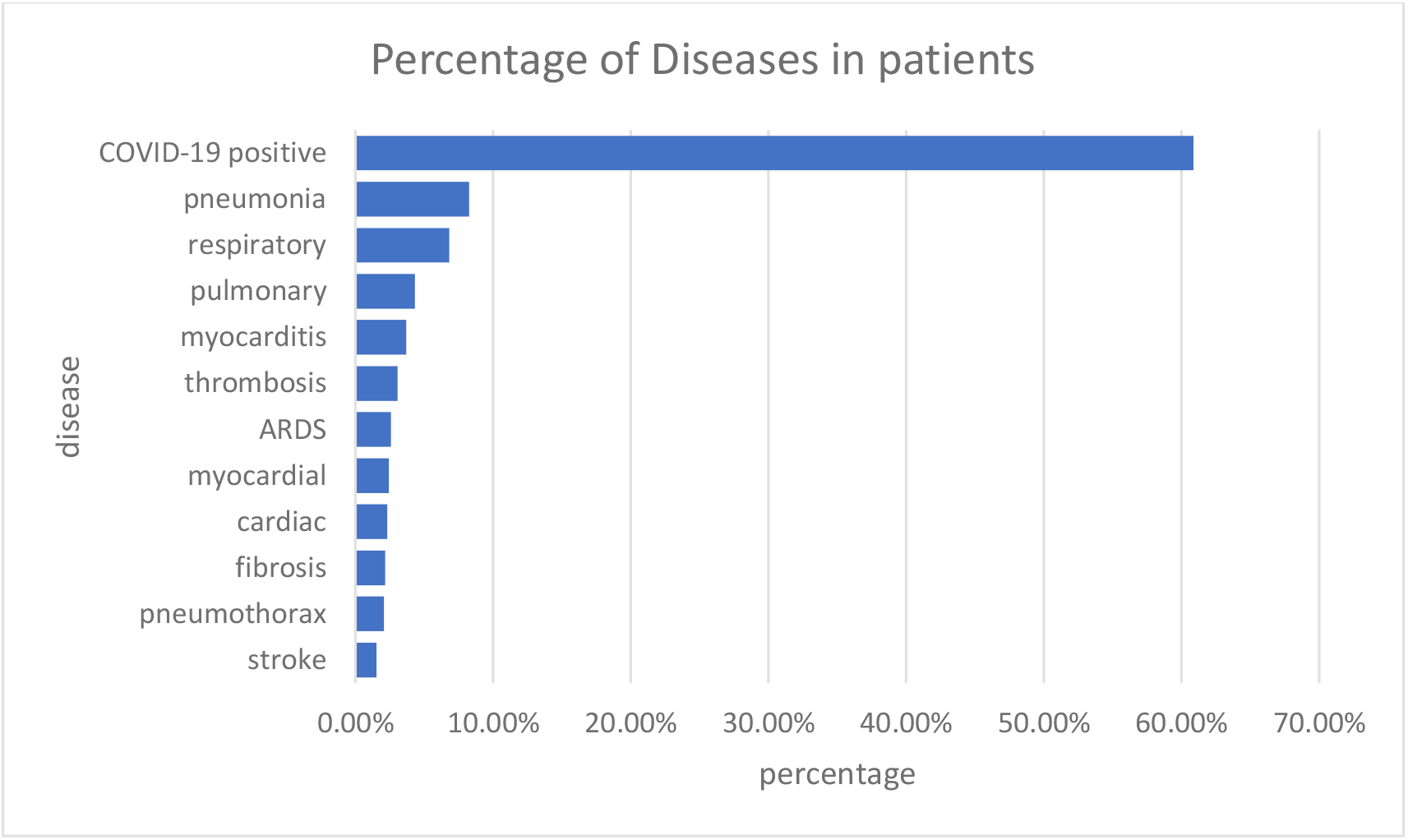
Percentage of most occurring disease disorders in patients.

We also show a snippet of one case report in PDF format that is parsed by our BioEN model and the results are saved and annotated in the same PDF file as shown in Figure 8.

**Figure 8:**
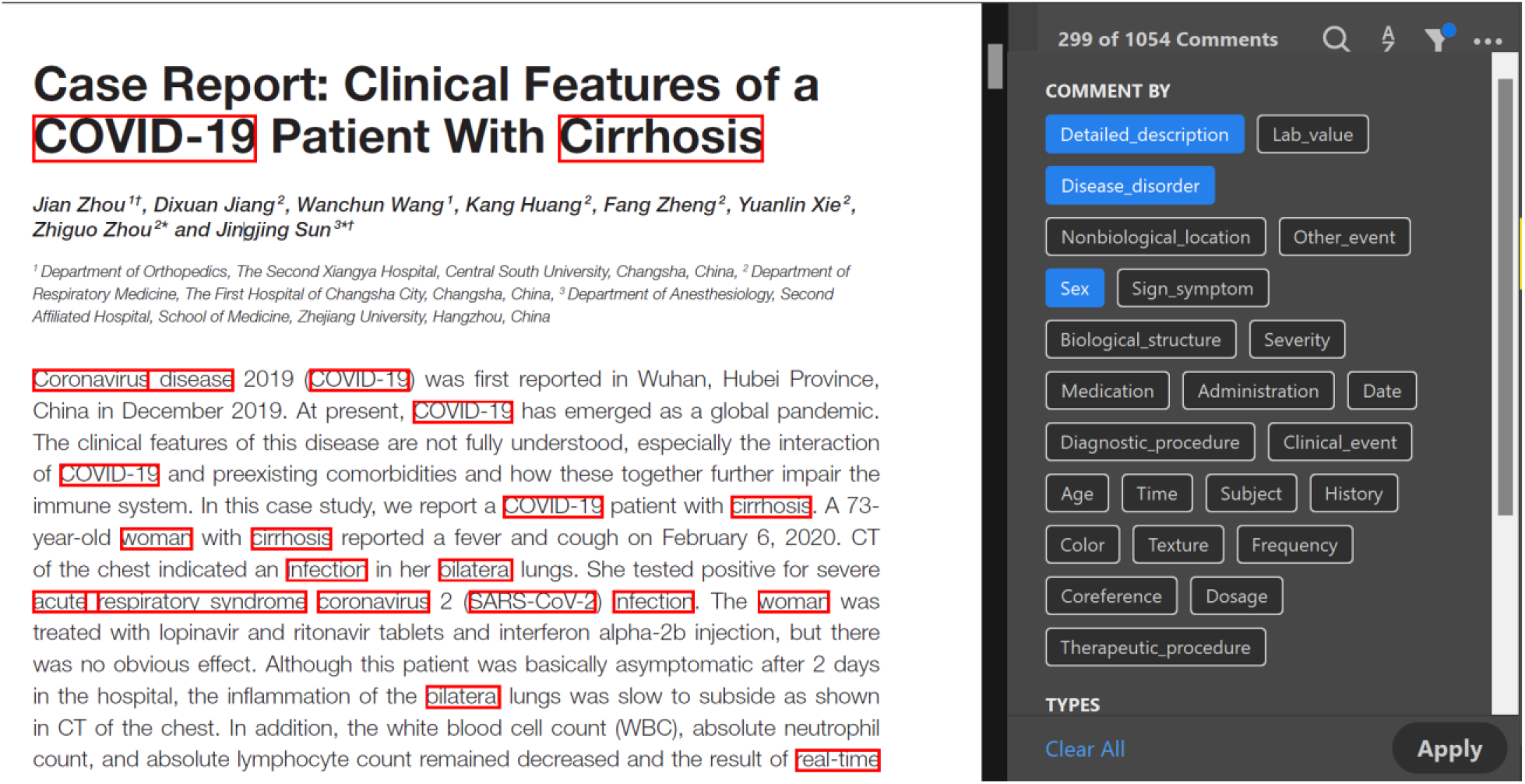
Annotations produced by our model in PDF file.

## 5 Discussions

This package’s results and findings can be used in healthcare applications, such as assisting doctors, nurses, and clinical experts in matching symptoms to diagnosis, treatment, and follow-up. This package is simple to install and configurable for real-time use by medical practitioners. The model’s results can be applied to a variety of applications, for example, to study the disease detection, demographic studies, social determinants of health, semantic relation extraction between concepts in medicine biology, and other related tasks. BioEN also has an impact on application performance in terms of precision and recall.

Healthcare and health science data face numerous challenges in the era of “big data”. With this approach, we attempt to provide automatic methods for text and data mining tools that must be deployed to deal with large, highly heterogeneous data sets. As the field NLP advances, policymakers will have more opportunities to understand the value contained in electronic medical records and clinical records, as well as the cost-effectiveness and cost-savings implications of health system planning. This solution also allows for the tracking of medical as well as social determinants of health, which can lead to the reduction of health disparities. More research, however, is required to better understand and assess the dataset.

### Limitations

So far, we rely on benchmark data to train the model. However, more data is required to train the model to study a large number of infectious diseases. In future, we plan to annotate our own biomedical data and we strongly encourage the inclusion of medical professionals in the annotation guideline. We also intend to curate more clinical data, in particular, getting real-time access to EHRs would be helpful. Due to the black-box nature of most deep neural networks, we also plan to handle bias or systematic error in research methods, which may influence disease associations and predictions. We also plan to consider the human evaluation to have predicted entities as being more accurate, informative and biomedical.

## 6 Conclusion

In conclusion, this paper presents BioEN development architecture that consists of a number of components stacked together. We use an approach to train models for the biomedical named entities using by fine-tuning the BERT based Transformer model. We fine-tune a Transformer based architecture to the task of biomedical NER. We evaluate the performance of our approach different benchmark datasets, and our approach achieves the state-of-the-art results compared to the baselines. We demonstrate through extensive experiments that using contextualized word embedding pre-trained on biomedical corpora significantly improves the results in NER task.

## Data Availability

https://figshare.com/articles/dataset/MACCROBAT2018/9764942

## Acknowledgement

We would like to acknowledge the Canadian Institutes of Health Research’s Institute of Health Services and Policy Research (CIHR-IHSPR) as part of the Equitable AI and Public Health cohort for supporting this research.

